# Cladribine tablets in Relapsing-Remitting Multiple Sclerosis preferentially target B-cells

**DOI:** 10.1101/2024.03.28.24304956

**Authors:** Francesca Ammoscato, Mohammad Aboulwafa, Justyna Skonieczna, Jonathan Bestwick, Rosemary Monero, Michael Andrews, Stefania De Trane, David Holden, Ashok Adams, Lucia Bianchi, Ben Turner, Monica Marta, Klaus Schmierer, David Baker, Gavin Giovannoni, Sharmilee Gnanapavan

## Abstract

Recently it has been shown that treatments targeting B cells in multiple sclerosis (MS) are effective in controlling disease activity. B cells contribute to the pathogenesis of MS via antigen presentation, T cell activation, and antibody production. In the chronic progressive cladribine trial, some patients treated with cladribine had a significant decline in oligoclonal band number. However, the mode of action of cladribine tablets (CladT) on peripheral immune cells and its biological activity within the CNS remains to be determined further.

The CladB study is a longitudinal prospective investigation of CladT treatment in relapsing-remitting MS (RRMS). Blood was sampled at Day 0, 1, 5, then once a week for 8 weeks, fortnightly up to 24 weeks, and once a month till 96 weeks for immune cells. This was compared to a historical cohort of alemtuzumab treated samples for one month. Paired cerebrospinal fluid (CSF) and blood were also taken at Day 0, 48 and 96 weeks after initiating CladT for Kappa and Lambda-free light chain (кFLC, λFLC) index, oligoclonal bands (OCBs), immunoglobulin indices, inflammatory mediators and neurofilament light chain (NfL). Participants also underwent clinical and magnetic resonance imaging brain assessments.

Ten participants (3 male, 7 female, mean age 35.9 ± 10.5 (SD) and Expanded disability Status Scale 2.5 (range 0-6) at baseline were enrolled. B cells, in particular memory B cells, were heavily depleted by CladT. Alemtuzumab, conversely rapidly depleted both T and B cells. Although still present, reduction in OCB numbers were observed in 4/10 participants and кFLC index reduced from mean 164.5 ± 227.1 (SD) at baseline to 71.3 ± 84.7 at 48 weeks (p=0.002) and 64.4 ± 67.3 at 96 weeks (p=0.01). This coincided with reduction in IgG index [1.1 ± 0.5 (SD) at baseline, 0.8 ± 0.4 (p=0.014) at 48 weeks and 0.8 ± 0.3 (P=0.02) at 96 weeks] and CSF CXCL-13 [88.6± 68.4 (SD) pg/mL, 39.4 ± 35.2 mg/mL (p=0.037) at 48 weeks and 19.1 ± 11.7pg/ml at 96 weeks (p=0.027)]. CSF NfL levels were reduced at 48 weeks only (p=0.01).

In conclusion, our study supports the view that CladT treatment works primarily by depleting memory B-cells and antibody-secreting cell precursors in RRMS leading to sustained effects on intrathecal antibody production and total IgG associated with a reduction in the B-cell chemoattractant CXCL-13 in the CSF.

## Introduction

MS is an autoimmune disorder and is invariably associated with an intrathecal oligoclonal IgG response or oligoclonal bands (OCBs)^1^. This response is not specific to MS and also occurs in central nervous system (CNS) infections and other CNS-specific autoimmune diseases, in particular the paraneoplastic syndromes^2^. In MS these OCBs may be targeting self-antigens, or maintain pro-inflammatory glial cell activity, and may therefore be responsible for driving progressive MS pathology, which theoretically could become independent of autoimmune T-cell help^1,3^. Pathological studies have shown that the presence of B-cell-like follicles in the meninges of people with MS (pwMS) are associated with earlier age of onset of progressive disease and earlier age of death^4^. Recent data has linked the subpial cortical MS lesion with antibody deposition and complement activation ^5,6^, similar to that which has been described in the so-called type 2 acute MS lesion ^7^.

We have recently hypothesised that all effective MS DMTs, particularly high-efficacy DMTs, work via targeting the peripheral memory B-cell population and suggested that cladribine tablets (CladT) may be preferentially working as a semi-selective B-cell agent^8^. We now have cross-sectional pilot data from pwMS treated with parenteral cladribine showing prolonged peripheral memory B-cell depletion of a similar magnitude to alemtuzumab-treated patients^8,9^. Moreover, CladT is a small molecule nucleoside analogue that has the potential to penetrate the blood-brain barrier leading to targeting of both peripheral and central inflammatory mechanisms. It may therefore exhibit activity against pathogenic B cells in the CNS and reduce intrathecal IgG and free-light chains with a potential disappearance in intrathecal OCBs. In a recent study, CladT demonstrated a reduction in memory B cells as early as one month after treatment initiation, with low levels sustained for up to 12 months^9^.

We therefore undertook an intensive sampling prospective longitudinal B-cell biomarker study (CLAD-B study) to explore the effect of CladT on peripheral and intrathecal B cells. We hypothesised that CladT preferentially targets the B cell population early, particularly the memory B cells, whilst the repopulation of B-cells is driven initially by naive and subsequently mature B-cells, but not memory B cells. Secondly, in people with MS (pwMS) the administration of CladT reduces CSF biomarkers of both B cell and plasma cell activity (OCBs and absolute free light chain levels) and markers of CNS inflammation (CXCL-13) and neuroaxonal damage (neurofilament light chain). Lastly, as with alemtuzumab CladT results in early depletion of the memory B-cell pool relative to the T cell compartment.

## Materials and Methods

### Study Design

CladB study was a prospective longitudinal biomarker investigation of CladT in Relapsing-remitting MS (RRMS). The study was approved by the Stanmore Research Ethics Committee (IRAS ID 262436; Rec ref 19/LO/0789).

Study participants (n=10) underwent blood sampling for peripheral blood mononuclear cells (PBMC) at the following time points: Day 0, 1, 5, then once a week for 8 weeks, fortnightly up to 24 weeks, and once a month till 96 weeks. Clinical scales (Expanded Disability Status Scale (EDSS), MS functional composite (MSFC), MS impact scale-29 (MSIS-29), Paced Auditory Serial Addition Test (PASAT). Nine-hole Peg test (9H-peg test (9H-PEG)) and MRI-Head were performed at Day 0, week 48 and week 96. CSF and urine were also collected at these time points.

Early changes in the first month in peripheral immune cells were compared with alemtuzumab treated participants (n=10) at week 1 Day 5 and week 4 (NCT06310343, unpublished work).

### Flow cytometry

#### Peripheral blood mononuclear cell (PBMC) preparation

PBMCs were obtained from blood by density gradient centrifugation using Lymphoprep and SepMate tubes (StemCell Technologies Vancouver, BC, Canada). Briefly, the blood was diluted in PBS (without Ca^2+^, Mg^2+^) and 25 ml of diluted blood was loaded onto 15 ml of Lymphoprep previously loaded in a SepMate tube. Tubes were centrifuged for 10 min at 1350 rpm at 20 °C. The buffy coat was collected into a 50 ml tube and washed twice in PBS. Finally, the pellet was resuspended in 5 ml PBS, cells were counted using a haemocytometer (Revvity Health Sciences, Inc.), resuspended in 1 mL of freezing medium (10 % dimethyl sulfoxide and 90 % fetal bovine serum) and transferred into cryovials. The tubes were then placed into a freezing container and stored at −80 °C freezer overnight. The frozen cryovials were transferred and placed in liquid nitrogen the day after.

#### PBMC recovery

Purified PBMCs were thawed by transferring them to a 37°C water bath and resuspended in 8 ml of RPMI 1640 medium (Gibco, Grand Island, NY, USA) at 37 °C. The suspension was then centrifuged for 10 minutes at 300 x g at room temperature. Thawed cells were washed and resuspended in 5 mL of pre-warmed complete RPMI. A 50 μL aliquot was removed for cell counting using a haemocytometer. After centrifugation, cells were resuspended in culture medium, distributed in wells of a 48-well flat-bottom plate (approximately 5-10 x 10^6^ PBMCs, 400 μl per well), and transferred to the incubator overnight at 37 °C. The following day, cells were stimulated with PMA (100 ng/ml) and Ionomycin (1 μg/ml).

#### Staining protocol

To determine the population percentages of lymphocyte subsets in peripheral blood mononuclear cells (PBMCs), a panel consisting of 18 fluorophores was designed. The panel (Supplementary materials) included markers: CD3, CD8, CD4, CD45RA, CCR7, CD25, CD127, and FoxP3 for fluorescent labelling of T cells, while CD19, CD20, CD27, IgD, CD38, CD138, CD24, CD10 and IL-10 were used to characterise B cells. Zombie NIR fixable viability dye was used to distinguish dead cells.

For sample preparation, tubes containing 1×10^6 cells were washed twice with staining buffer (1% BSA in PBS), centrifuged at 300g for 5 minutes at 4°C, and resuspended in a final concentration of 100 µl of the master mix containing 1:1000 Zombie NIR, Fc Blocking (Thermofisher, Waltham, Massachusetts) and staining buffer. After a 30-minute incubation at 4°C, cells were washed and resuspended to a final volume of 100 µL with the antibody master mix (Supplementary materials), Brilliant stain buffer (BD, Bioscience, New Jersey, USA), and incubated for 30 minutes at room temperature, protected from light. Subsequently, stained cell samples were washed twice by adding 500 µL of staining buffer to each sample and centrifuged at 300 x g for 5 minutes. After the wash, cells were fixed with Foxp3 Fixation/Permeabilization (eBiosceince, Waltham, Massachusetts) working solution, left on ice for 30 minutes, and then washed with Flow Cytometry Perm Buffer. After centrifugation, cells were stained with antibodies against intracellular antigens (FOXP3 and IL-10) and incubated for 1 hour at room temperature. The cells were then washed with 1 ml of Perm buffer, resuspended in staining buffer, and were ready to be acquired on a Cytek Aurora. The results for IL-10 are not presented in this study as no differences were observed in IL-10^+^ levels across all the time points.

### Body Fluid Biomarkers

#### Kappa and lambda free light chain levels and index

Serum and cell free CSF samples from patients were stored in -80°C until the analysis. Kappa Free Light Chains (кFLC, LK016.OPT), Lambda Free Light Chains (λFLC, LK018.OPT), IgG (NK004.LL.OPT), IgM (LK012.L.OPT), IgA (LK010.L.OPT) and albumin (Alb, NK032.L.OPT) were measured on the Optilite™ turbidimetric analyser (The Binding Site®, Birmingham, UK) with kits supplied by the producer. The samples were not manually prediluted and inputted directly into the analyser after thawing. The analyser automatically recorded kinetics of the reaction to control and checked the antigen excess in samples to automatically adjust dilutions if required. The dilutions used for the assays were as follows: for CSF samples 1:1 in all assays, for serum samples 1:8 up to 1:10 for кFLC and λFLC, respectively; 1:199 for IgG, 1:399 for IgM, 1:799 for IgA and 1:200 for Alb. All assays were conducted as per the Optilite analyser manual. Index calculations were performed as follows: кFLC index=Q_κFLC_/Q_Alb_, λFLC index=Q_λFLC_/Q_Alb_, IgG index=Q_IgG_/Q_Alb_, IgA index=Q_IgA_/Q_Alb_, IgM index=Q_IgM_/Q_Alb_.

#### CSF and serum Oligoclonal Bands

CSF and matching serum samples from patients were stored at -80°C until analysis. IgG oligoclonal bands were analysed by isoelectric focusing (IEF) on the Multiphor II electrophoresis unit (Pharmacia Ltd.). IEF gels were prepared in-house, stored at 4°C, but brought to room temperature before analysis. The serum samples were manually prediluted 1:400, but the CSF were run neat. After IEF, the samples were immunoblotted onto a nitrocellulose membrane. The membrane was blocked in non-fat milk and then probed overnight with 1:1000 dilution Anti-human IgG (Sigma). The following day, the membrane was washed then probed with a 1:1000 dilution Goat anti-rabbit IgG HRP. It was washed again and visualised with ethyl-amino-Carbazole. The Oligoclonal bands in both serum and CSF were interpreted by the analyst then second checked and authorised by senior staff. Gel band quantification was performed on ImageJ.

#### CSF neurofilament light chain levels

Cell free CSF were stored at -80 °C until analysis. NfL was analysed using the SiMoA NF-light Advantage Kit (Quanterix, Bellerica, MA, USA) on the Simoa HD-X bead-based immunoassay analyzer (Quanterix, Billerica, MA) as per protocol. Briefly, CSF, calibrators, and controls were thawed at room temperature and directly transferred, undiluted, to Quanterix microtiter plates. The plates were sealed to prevent evaporation. Kit components (bead reagent, detector reagent, streptavidin-ß-galactosidase (SBG) reagent, resorufin ß-D-galactopyranoside (RGP) reagent) were barcode-scanned before placement in the instrument. The plate set-up also involved configuring the CSF dilution at a ratio of 1:100. Post-assay, Quanterix software facilitated data analysis. Calibration signals were fitted to a 1/Y2-weighted four-parameter logistic (4PL) curve, employed to calculate NfL concentrations in samples.

#### CSF cytokines, CXCL13, complement and sCD138

Th1 and Th2 (IFN-γ, IL-1β, IL-2, IL-4, IL-5, IL-8, IL-10, IL12p70, IL-13, TNF-α) and human BCA1/BCL were measured using Mesoscale Discovery (MSD) platform in the CSF; U-plex Th1/Th2 markers (K15010K) and R-plex for BCA1/BCL (Assay (1) from the kits protocol) (K1519QR). The dilution used for samples was 1:1 in the corresponding kits diluents. Syndecan-1/CD138 with R-plex MSD assay (1) from the kits protocol (K151V4R) used 1:2 dilution. Complement C3 R-plex MSD assay (K151XYR-2) used 1:1 dilution.

### MRI analysis

All patients underwent MR imaging with Siemens Magnetom Avanto_fit 1.5 Tesla MRI scanner. All the patients underwent three MRI examinations: baseline before starting Cladribine tablets, year one post therapy and year two post therapy. The MRI head protocol included an unenhanced T1 MPRAGE sequence [Voxel size: 1.0×1.0×1.0 i.e. isotropic voxels], sagittal 3D FLAIR sequence [Voxel size 0.5×0.5×1.0], sagittal 3D DIR [Voxel size: 1.5×1.5×1.5] and an axial post-contrast T1-weighted sequence. Two experienced raters (AA, neuroradiologist, and MA, trained neurologist) independently evaluated lesion activity on different time points blinded to patient clinical data. Disease activity was reported by the number of new, enlarging, or contrast-enhancing lesions compared to the corresponding previous time point.

### Statistics

The primary endpoint was to look at the effect of CladT on peripheral immunological markers over time. However, to explore the intrathecal effect of CladT we based the sample size calculations on the likelihood of disappearance of OCBs from the CSF. Based on published data none of the patients with MS will be expected to lose OCBs over 2 years of the study^10,11^.

Using a one-way pairwise ANOVA with a power of 80%, and an alpha of 5%, we will only need 10 subjects to show a significant result if at least 50% of subjects on cladribine become OCB negative compared to a comparator group. Allowing for a 20% dropout rate we would only need to recruit 12 subjects. However, owing to the changes in the management of MS during the COVID-19 pandemic and subsequent under recruitment we closed the study at 10 subjects.

Data were described using percentages (categorical variables) or means and standard deviations or medians and ranges as appropriate (continuous variables). Comparisons between B and T cells at specified time points were made using Wilcoxon signed rank tests. Associations between percentage change in T and B cells and clinical parameters over the course of the study were described using Spearman’s rank correlation coefficients. Statistical significance was taken as p<0.05. Given the sample size, no adjustments were made for multiple testing. All analyses were performed using Stata v18 (StatCorp, College Station, Texas).

## Results

### Patient characteristics

Out of twelve, ten participants completed 96 weeks in the CladB study and were included in the final analysis (Table 1). The cohort were predominantly young female adults, with established MS, but low disability levels (median EDSS 2.5) excluding one individual and started CladT as their second line DMT. Six participants had other medical illnesses including osteoarthritis, hysterectomy, Raynauds phenomenon, asthma, systemic lupus erythematosus (on hydroxychloroquine), hypothyroidism, hypercholesterolemia, asthma, anterior uveitis, and hidroadenitis suppurativa. Regarding disease activity participants had experienced on average 2.0 relapses since diagnosis, half of whom had evidence of Gadolinium (Gd) enhancing lesions with 80% demonstrating new/enlarging T2 lesion activity (range 1-4).

**Table 1.** Demographic and clinical characteristics of study participants.

| Characteristic |  | N(%) / Mean(SD) |
| --- | --- | --- |
| Sex | Male | 3 (30%) |
|  | Female | 7 (70%) |
| Age |  | 35.9 (10.5) |
| Ethnicity | Caucasian | 5 (50%) |
|  | Black | 4 (40%) |
|  | Asian | 1 (10%) |
| Comorbidities |  | 6 (60%) |
| Disease duration<sup>a</sup> |  | 56.3 (98.1) |
| Prior relapses<sup>β</sup> |  | 2.0 (1-11) |
| Prior DMT | None | 3 (30%) |
|  | Interferon beta | 2 (20%) |
|  | Tecfidera | 5 (50%) |
| Baseline EDSS<sup>β</sup> |  | 2.5 (0-6) |
| New Gd+ MRI lesion last 1 year |  | 5 (50%) |
| New/enlarging MRI T2 lesions last 1 year |  | 17 (80%) |
| OCB positive |  | 9 (90%) |
| Baseline PEG Test |  |  |
| Dominant hand, seconds |  | 23.76 (5.8) |
| Non-dominant hand, seconds |  | 23.71 (7.1) |
| Baseline T25 Foot Walk, seconds |  | 5.11 (1.78) |
| Baseline PASAT |  | 39.22 (0.9) |
| Baseline MSIS |  | 53.2 (15.6) |

<sup>a</sup> Average disease duration reported in months. <sup>β</sup> Median and range. SD=standard deviation

### Temporal changes in peripheral T cells after CladT

The effect of CladT on T cells is less dramatic compared to that on B cells (Figure 2), following an initial suboptimal depletion there is an elevation in numbers of T cells noticeable at Day 5 (72.5% CD3+CD4+ p=0.059, 50.8% CD3+CD8+ p=0.093), with the greatest rise in central memory T cells (68.6% CD4+, 174.9% CD8+ [CD45-CCR7+], p=0.047 and p=0.047, respectively) and naive T cells (107.2% CD4+, 72.5% CD8+ [CD45+CCR7+] p=0.037 and p=0.074, respectively). Thereafter, their levels drop and rise in the first year, but the greatest depletion in numbers is evident after the second course of CladT at week 48, after which T cells reach their nadir levels between week 60 (−81.1% CD3+, CD4+ p=0.028) and week 72 (−74.2% CD3+, CD8+ p=0.069). Both the central memory and naive T cells are depleted more effectively during the second round leading to less recapitulation of events that took place after the initial course. Similarly, the effect of CladT on depleting the CD4/CD8 effector memory (CD45RA-CCR7-) or TEMRA (CD45+CCR7-) population after the first course of CladT is less efficacious than after the second course; with levels again reaching their nadir around week 60-72. Early rises in regulatory FoxP3+ cells were evident week 3 (119.3% p=0.038) and intermittently thereafter, but like all other cells depleted again after the second course of CladT.

**Figure 1.**
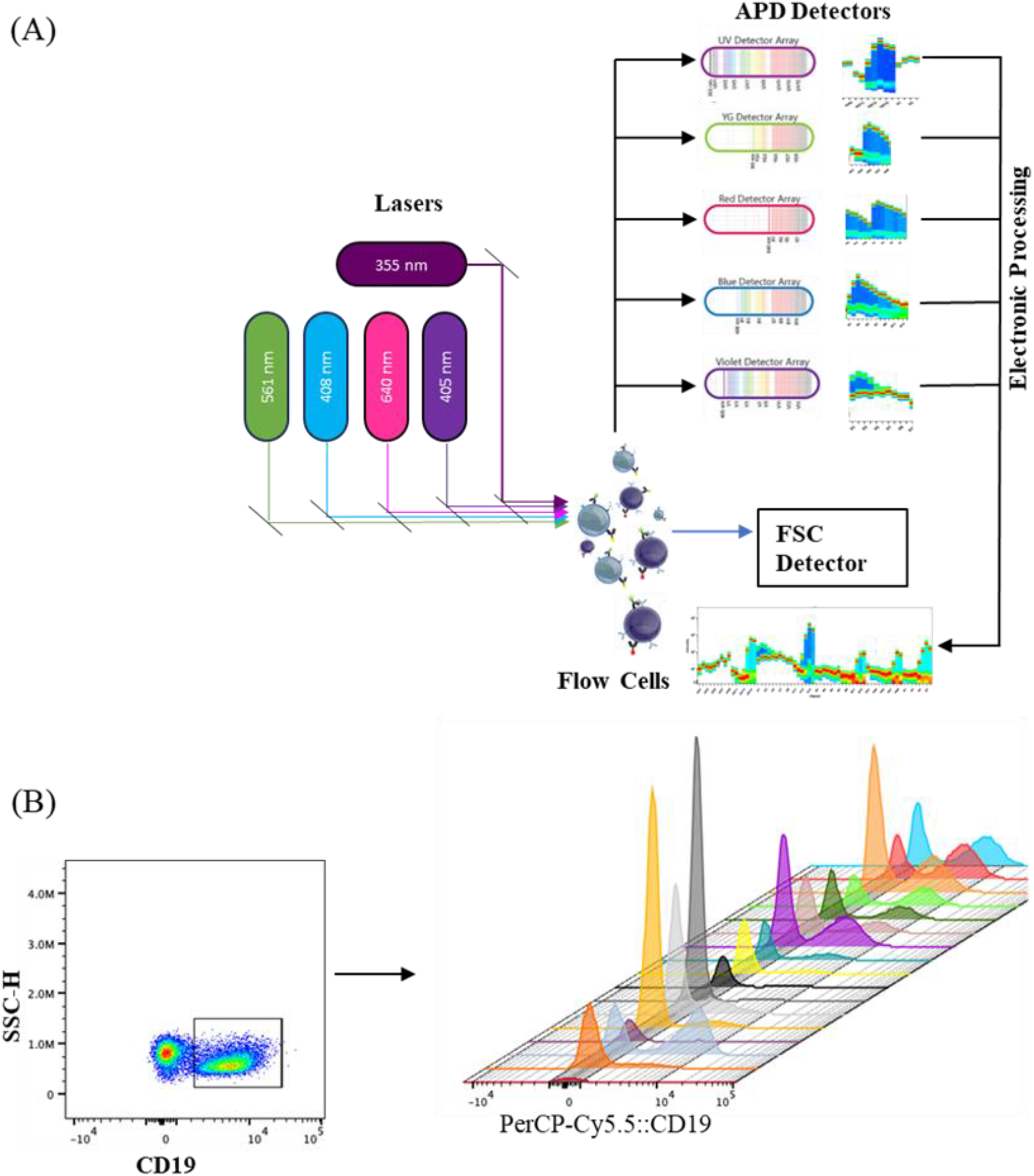
Full Spectral Flow Cytometry Diagram (A) The mechanism involves an optical setup with specialised filters and detectors that capture the emitted fluorescence. The captured signals from each detector are then combined, amplified and digitised to form a unified spectrum. (B) Representative dot plot and Histogram of CD19-PerCpCY5.5, from one of the participants, at baseline, visit1d1. v1d5, v2, V3, V4, V5..V6,V7,V8 etc…until visit 24

**Figure 2.**
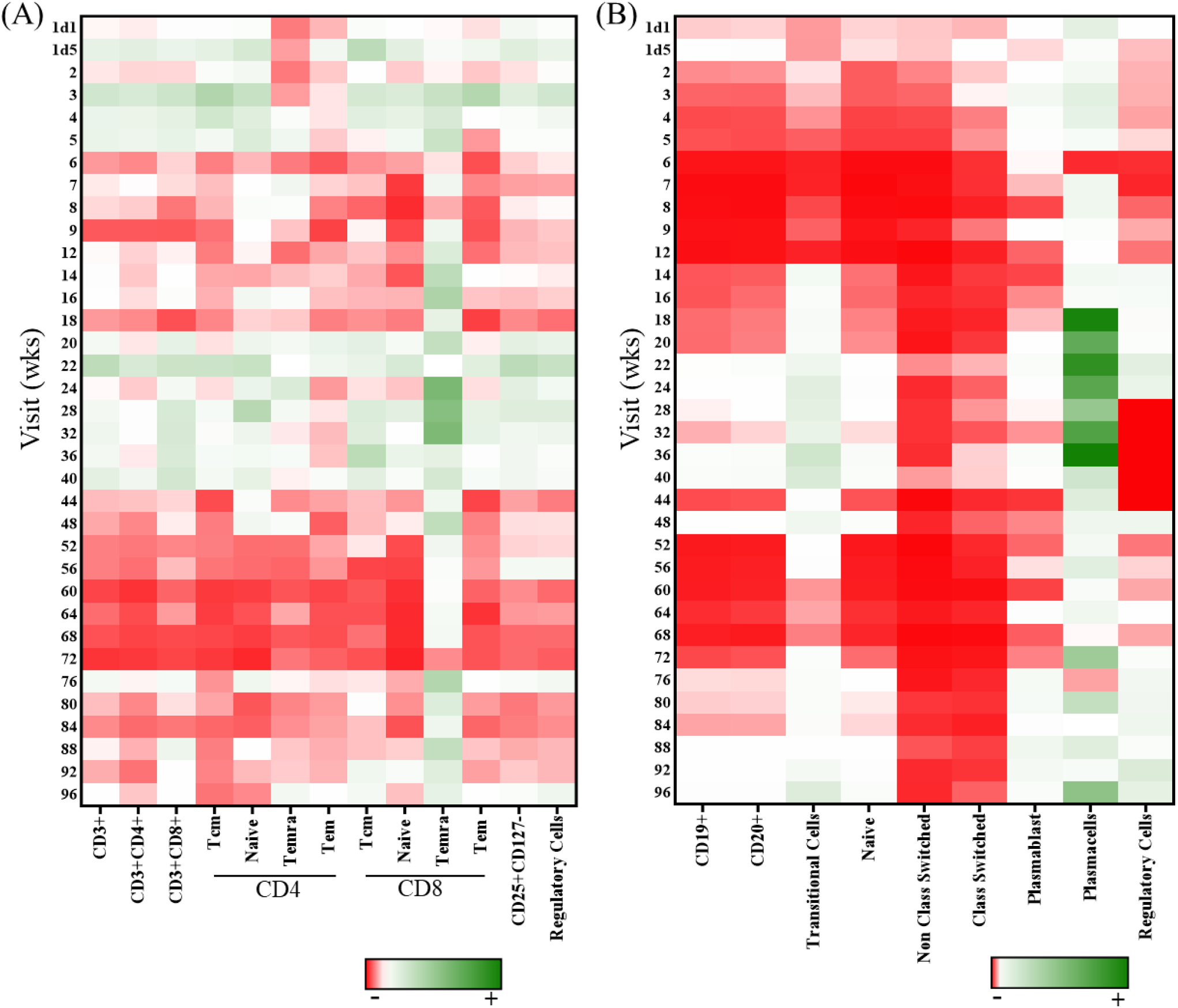
Heatmap showing data corresponding to the percentage change from baseline value for each subset at each timepoint. The expression intensity is displayed on a scale from red (negative) to green (positive) (A) T cell population (B) B cell population

### Temporal changes in peripheral B cells, plasmablasts and plasma cells after CladT

The effect of CladT depletion is a lot more dramatic on B cells (CD19+/20+) than in any other cell type, with prominent depletion in the memory B-cell pool (class-switched and non-class switched, Figure 2) evident within Day 1 of treatment but extending throughout the course of treatment. The non-class switched memory B-cells (CD27+, IgD+) were depleted more in terms of relative amounts than class-switched memory B-cells (CD27+, IgD-), but both reach nadir levels by week 6 (−96.4% p=0.005 and -82.6% p=0.007, respectively). The extent of depletion was similarly sustained at very low levels for a protracted period; until week 36 in the non-class switched population *vs.* week 20 in the class-switched population following the first course of CladT. This trend was maintained to the end of the study with the non-class switched population affected more (−85.3%, p=0.114) than the class-switched memory B-cells (−62.8%, p=0.203)

Similar to the memory B cells, the naive B-cells from the bone marrow (IgD+, CD27) are also heavily depleted by CladT reaching nadir levels at week 6 (−96.3% p=0.005) but recovering by week 14. Whilst the transitional B-cell population (IgD+, CD10+, CD27-) from the bone marrow demonstrate some of the earliest depletion (Day 1 -40.3% p=0.441) reaching again nadir levels at week 6 (−88.0% p=0.007). But, unlike the other cell types discussed earlier, this population demonstrates a resurgence in numbers at week 14 (288.1% p=0.051) till week 48 (374.1% p=0.037) when the second course of CladT is taken. As a result of repopulation, the effect of the second course of CladT is less evident and/or sustained with depletion taking place at week 60 (−44.7% p=0.600) to 72 (−50.7% p=0.093), and a second resurgence leading to peak numbers by 96 weeks (858.6% p=0.007). By comparison, naive B cells also start their recovery but at a later time point (week 22) following the first course of CladT and as such are again rapidly depleted after the second course of CladT to their lowest levels at 90.3% (p=0.017) by week 56. Their recovery, however, starts at the same time point (week 76) as the transitional B cells reaching a high of 136.6% (p=0.037) at week 96 after the second course of CladT. The regulatory B cell pool respond in a similar way to CladT as they fluctuate up and down in numbers with the initial resurgence happening at week 14 (244.2% p=0.051) with a less effective depletion after the second course resulting in a repletion of 648.3% (p=0.022) by week 96.

Plasmablast numbers increase from Day 1 of CladT treatment reaching high numbers at week 3 (309.1% p=0.036) but deplete afterwards with the second depletion course of CladT. Interestingly, their depletion seems to follow that of memory and naive B cells reaching peak depletion at week 8 (−73.5% p=0.953) following the first cycle of CladT and week 60 (−75.0% p=0.917) following the second cycle of CladT. Plasma cells (CD138^+^) on the other hand were largely unaffected by CladT, except at week 6 (−84.5% p=0.859) when plasma cell numbers suddenly dropped, coinciding with peak depletion of plasmablasts. Their numbers increase in both instances to peak levels during periods where other B cells and plasmablasts are showing their greatest depletion.

### CladT *versus* alemtuzumab in early peripheral cell depletion

Whereas CladT predominantly depletes B-cells, alemtuzumab acts more comprehensively on both T- and B-cells including the effector population (Table 2). As such only the plasma cell population boosted their numbers by week 4 (CladT 606.7% *vs* alemtuzumab 1585.0% p=0.355). The naive T-cells from the bone marrow were most susceptible to alemtuzumab depletion, particularly the CD4+ population (Day -79.83%), although by week 4 the CD8+ cells also deplete (−99.93%). But, similar to CladT, the non-class switched memory B-cells (CD27+IgD+ -100%) were heavily depleted by alemtuzumab also.

**Table 2.**
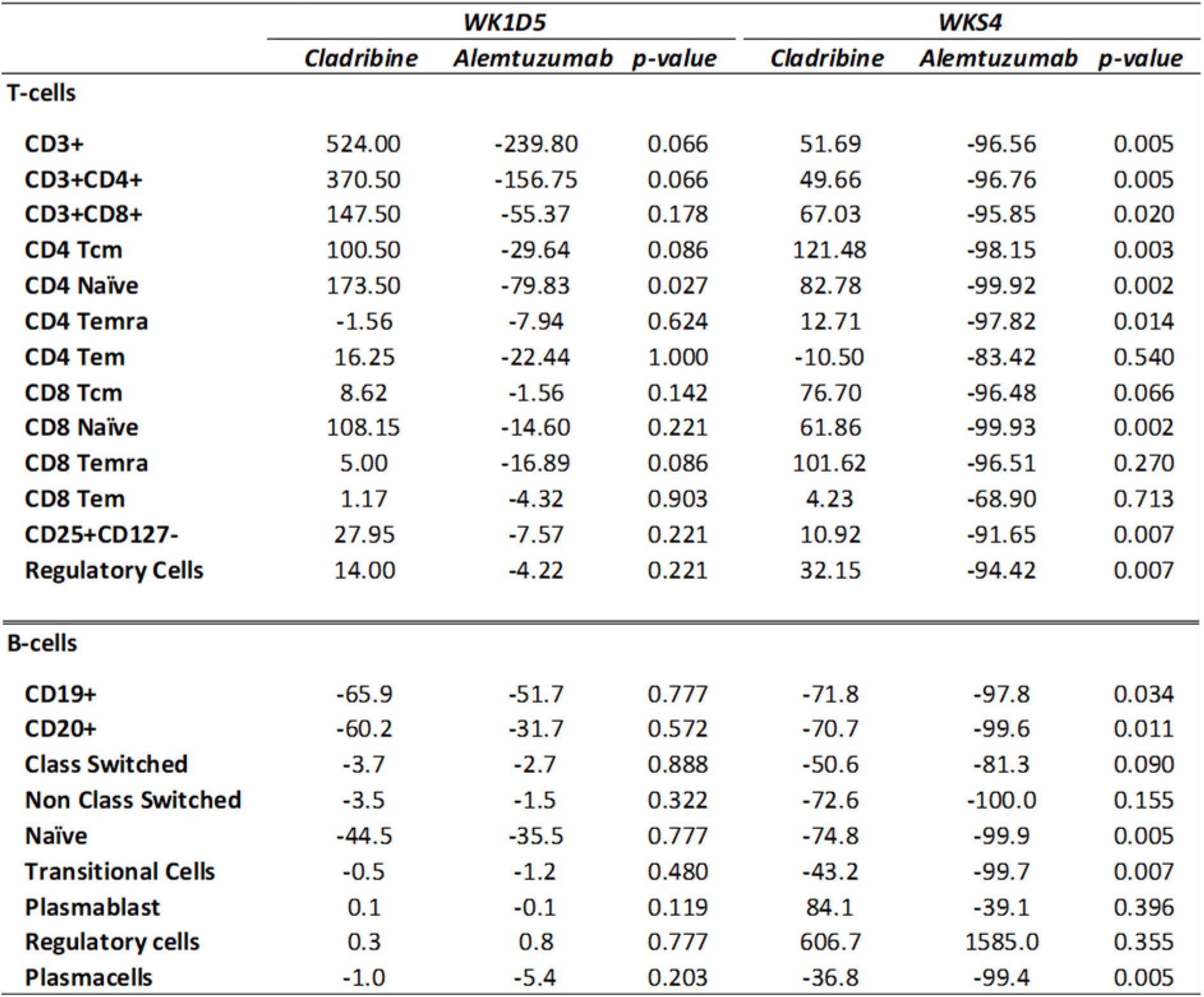
CladT vs. Alemtuzumab median % change in baseline cell numbers in the first month of treatment.

### Biological effects of CladT in the CSF

#### Oligoclonal bands and densitometry

In our cohort between baseline, week 48 and week 96, there was no individual that became oligoclonal band negative (Supplementary Figure 5). However, by densitometry evaluation 4/10 individuals had a reduction in band numbers, whilst 2/10 remained unchanged and increased in 2/10, while initially reducing at week 48 but increasing again at week 96 in 1/10. Unexpectedly, one individual was OCB negative at the start of the study and remained negative throughout. We found that OCB interpretation by visual inspection did not always conform to band densitometry assessments.

#### CSF free light chain levels, pro-inflammatory biomarkers and NfL levels

Up to week 96 there was a sustained reduction in CSF-кFLC levels but not λFLC levels (p=0.01) following CladT treatment (Figure 3). Coinciding with this reduction there is also a reduction in intrathecal IgG production (IgG index p=0.02) and B-cell chemoattractant CXCL-13 levels in the CSF (Figure 3, p=0.027), whilst peripheral immunoglobulin levels remain stable (data not shown).

**Figure 3.**
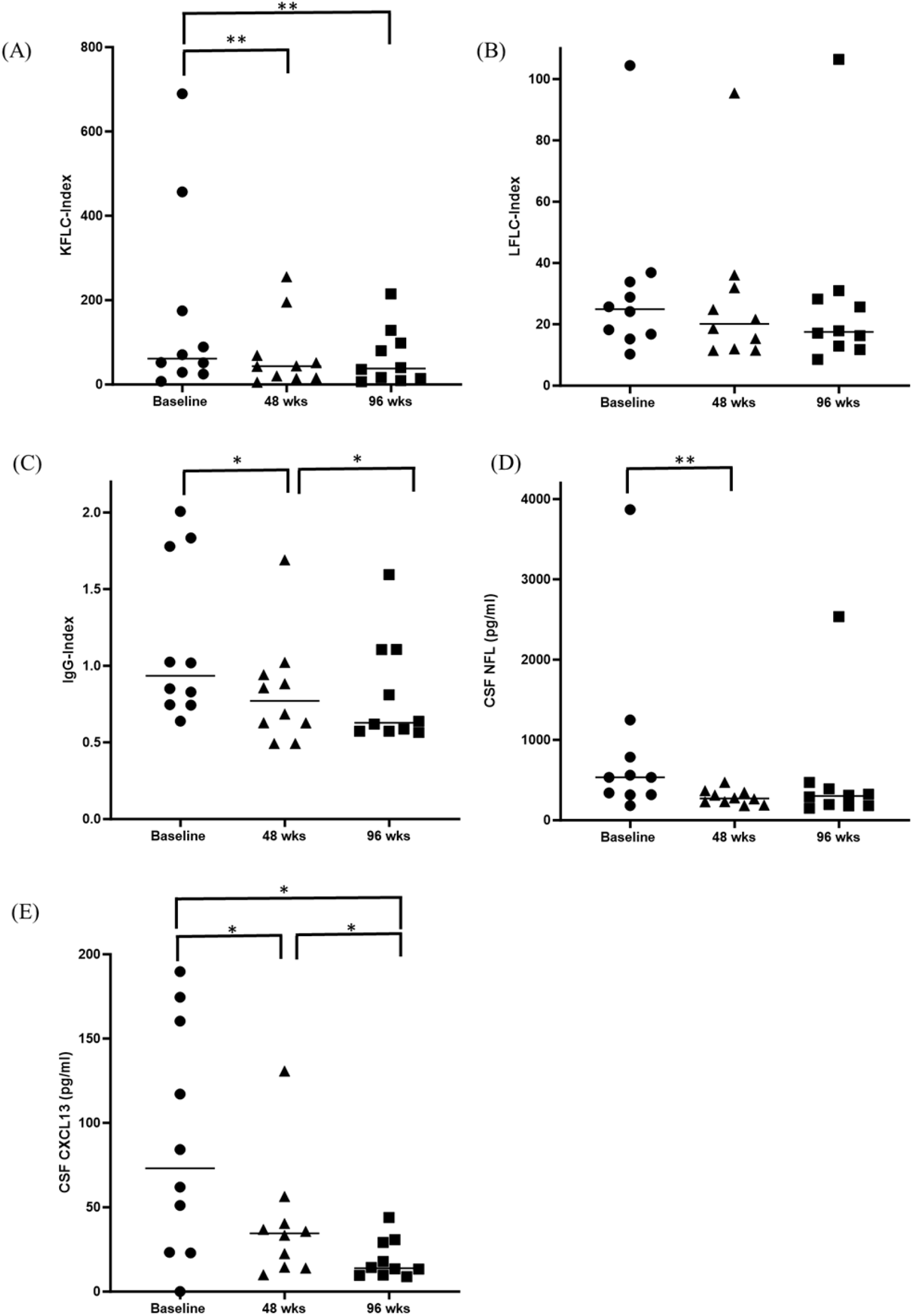
Biological effects of CladT on biomarkers: (A) Kappa free light chain Index (кFLC) (B) Lambda free light chain (λFLC) index (C) IgG Index in CSF/serum at baseline, week 48 and week 96 (D) Neurofilaments light chain (NFL) levels and (E) C-X-C motif chemokine ligand 13 (CXCL13) levels in CSF at baseline, week 48 and week 96.

CSF Th1/2 T-cell proinflammatory cytokines remained unchanged over the course of the study, and similarly other inflammatory biomarkers such as CSF complement 3c and urinary neopterin levels. CSF soluble CD138, a marker of plasma cells, was also unchanged (Supplementary Table 3).

Following the first cycle of CladT there was a significant reduction in CSF-NfL levels at week 48 (p=0.01), but this was not sustained to week 96 (Figure 3).

### Clinical effects of CladT based on changes in peripheral immune cells

Rank correlations of percentage change in T and B cells over the course of the study (Supplementary Table 5) were mainly evident between T cells and 9H-PEG test (dominant, DH). Percentage increase in T-cells from baseline to 48 weeks were associated with worsening 9H-PEG DH for increases in CD8+ T cells (*r=*0.714, p=0.05), TEMRA T-cells (*r*=0.857, p=0.010), Tregs (*r=*0.762, p=0.032), FoxP3 Tregs (*r*=0.738, p=0.040) and plasmablasts (*r*=0.881, p=0.007). Later changes in cells from baseline to 96 weeks were not associated with changes in the 9H-PEG test. Early changes in new MRI T2 lesions from baseline to week 48 similarly were associated with increased CD8+ T cells (*r=*0.674, p=0.034) and TEMRA T cells (*r*=0.652, p=0.043). Later changes in CD8+ T cells (*r*=-0.821, p=0.029) and naive T cells (*r=*-0.821, p=0.029) were observed with the PASAT.

## Discussion

In this prospective, longitudinal study of CladT in RRMS we demonstrate for the first time a reduction in intrathecal antibody production in the form of кFLC following its selective action on lymphocytes, in particular B cells. We also noted a reduction in CXCL-13, a B cell chemoattractant, and IgG index within the CSF whereas peripheral immunoglobulin levels (IgG, IgM and IgA) remain unaffected^12–15^. The impact of CladT on intrathecal immunoglobulin production has not previously been demonstrated and the majority if not all immunotherapies currently available in MS target the peripheral immune system. CladT is CNS penetrant reaching the CSF at 25% levels of the plasma concentration^16^ and can potentially reduce the number of lymphocytes that have been recruited into the CNS^17^ and have a direct impact on intrathecal antibody production.

The efficacy and safety of oral cladribine has been demonstrated in CLARITY^18^ and CLARITY EXT^19^. CladT is a purine analogue that accumulates in cells such as lymphocytes that have high ratio of the enzyme deoxycytidine kinase (DCK)/5’ nucleotidase that activate and deactivate CladT metabolism, respectively leading to accumulation within cells and cytotoxicity^20,21^. Our analysis shows a greater impact of CladT on B cells compared to T cells; and also a greater impact on non class-switched versus class-switched memory B cells. This depletion occurs as early as Day 1 from the start of treatment reaching nadir levels by week 6 and is then sustained at least to week 96. Conversely within the T cells population there is an early compensatory rise in central memory (from lymphoid organs) and naive T cells (from the bone marrow) at Day 5 maintaining the body’s adaptive defence despite the loss of mature B cells. This homeostatic proliferation of memory T cells was seen with alemtuzumab also ^22^. Similarly, there was a compensatory rise from plasmablasts and plasma cells. This differential susceptibility may be secondary to in dCK and 5’ nucleotidase expression and/or activity in the two cell populations ^23,24^. As such it has been demonstrated that the dCK activity is significantly lower in unstimulated T cells compared to stimulated T cells exposed to CladT^20,23^, including differences in CD4 and CD8 T cell subsets ^23,24^. Both dCK and 5’nucleotidases are similar in naïve and memory B cells ^23,24^. CladT significantly reduces the number of clones and clonal diversity in memory B cells and as a result the potential for clonal expansion^25^. However, dCK expression is lower in plasma cells, this would be consistent with a more limited depleting effect on CD138+ plasma cells ^26^.

The time course of cellular depletion/repletion following CladT treatment varies between studies depending on the frequency of blood monitoring. Since our study has been the most intensive to date, we were able to note the early depletion in memory B cells and a compensatory response from T cells. In other peripheral blood immune cell profiling studies on oral cladribine a persistent decrease in B-cells, particularly in memory B cells has been documented by week 4 following the first treatment course^13–15,27^. Whereas the evidence on the effect of CladT on T cells is more variable with some studies noting a significant decrease in T cell number with delayed repletion^15,28^, whilst others reported a less pronounced effect on the overall T cell population with involvement of CD4+ T cells or central memory T cells^26,29,30^, or no or limited effect on CD8+ T cells^27,29^. We noted in our cohort better depletion in CD4+/CD8+ central memory, effector memory and TEMRA T cells after the second cycle of CladT ^26^, but with an early compensatory rise regulatory T cells (FoxP3+) after the first course of CladT reaching a high at week 3 similar to some other findings^31^.

Recovery of B cells numbers was first noted in our study at week 14 in the naive and transitional B cell populations that have potentially recently left the bone marrow. Despite this the memory B cell pool (non-class switched predominantly) remains depleted to week 96. Regulatory B cells also demonstrate early recovery at week 14, but unlike their T cell counterparts do not deplete effectively following the second cycle of CladT ^26^. It is not known what the significance of these differences between the two populations are. Regulatory B-cells are thought to exhibit T cell regulatory activity, notably because they produce interleukin 10 ^32^, they will also contribute to re-establishing normal B cell balance, as IL-10 also promotes activation and differentiation of B cells and plasma cells ^33,34^. As such, early repopulation of the peripheral blood with regulatory B cells is a stereotyped response seen with other B cell depleting agents^35^.. Plasmablasts unlike memory B cells rise early at week 3, although like the other cell types they also deplete at similar time points. Whether plasmablasts are susceptible to the effects of CladT after migration from the spleen/lymph nodes into the peripheral circulation or the exposure is taking place in the secondary lymphoid organs is unknown. However, their early rise in numbers suggest this is a compensatory response and unlike our hypothesis this equilibration takes place in the B cell population before the demand for more cells is met by cells from the bone marrow, such as the naive and transitional B-cells.

Plasma cells are relatively unaffected by CladT consistent with their more limited expression of dCK ^7^ and increase in their overall population size over the two treatment cycles and this may also be a compensatory response to the significant depletion in circulating B cells or may help serve to limit infection during lymphocyte depletion. This together with the early repletion of naive and transitional B cells may explain why patients on CladT are able to demonstrate and maintain a vaccine response with COVID-19 vaccines^36,37^. The lack of activity on plasma cells may also explain why in our cohort we did not observe anyone becoming OCB negative, whilst the reduction in band numbers in 4 out of 10 patients by densitometry suggests as with reduction in кFLC and IgG index that some of the antibody secreting cells originate from circulating effector B cells depleted by CladT. In this study we did not have data to assess the long-term effects of CladT but in a 10 year follow up study of individuals receiving subcutaneous cladribine 55% did not have CSF OCBs compared to baseline^38^, whilst in a chronic progressive cladribine trial, patients treated with cladribine had a significant decline only in the number of OCBs^39^. The evaluation of OCBs is very rater-dependent and may also explain the variable results observed across the studies. Over the last decade кFLC have emerged as a new quantitative biomarker of intrathecal antibody production that is rater-independent unlike the OCBs with the same diagnostic accuracy in MS as OCBs^40^. Although ours is the first study to evaluate this with CladT treatment.

Comparisons of percentage change in absolute numbers of peripheral cells in the short-term *vs*. long-term were mainly changes in the peripheral T cell compartment as opposed to B cells and mainly with the PEG (dominant hand) early on (up to week 48), but also with new T2 lesions. This despite the largest change being observed in the memory B population following CladT. However, this data set is small and interpretation of these observations are exploratory in nature and viewed with caution.

Comparison of early changes of CladT and alemtuzumab induction demonstrate very different peripheral immune cell depletion kinetics. The speed and completeness of depletion of both T and B cells by alemtuzumab by week 4 is very different to that of CladT resulting in immunosuppression. But, similar to CladT, the non-class switched memory B cells are most susceptible to depletion and plasma cells remain unaffected. Cencioni et al. also point to specific targeting of memory B cells by alemtuzumab, whilst Baker et al. also found T cell depletion:; 70-95% CD4+ and 47-55% CD8+ T cell depletion^30,41^.

CladT treatment resulted in a significant reduction in CSF NfL levels at week 48 but this was not sustained up to week 96. CladT is administered in RRMS as an induction treatment consisting of 2 treatment courses a year apart, and it is possible that there is a wearing off effect by week 96 and there was indeed some disease breakthrough in our cohort as would be expected based on No-Evidence of Disease Activity within the CLARITY studies ^18^.

There are no published studies that determine the biological efficacy of CladT on CSF-NfL levels. In a recent analysis of the MAGNIFY study, which examines a larger cohort of pwMS^9^ data serum NfL (sNfL) levels nonetheless did demonstrate a reduction in median sNfL Z-scores at month 12 and 24 ^42^. Whilst a much smaller analysis consisting of only 14 patients, demonstrated that the majority showed significant reductions in sNfL at month 12 barring one patient where the sNfL increased^43^. A third course of oral cladribine, however, is only available for breakthrough disease activity beyond year 4^44^.

We did not note any changes in proinflammatory cytokine or chemokine profiles in the CSF following oral cladribine, although others have noted a reduction in IL-2, sIL-2R, IL-8 and RANTES after treatment^45,46^. Neither did we note any changes in complement after oral cladribine treatment. Historically, one study has looked at the effect of subcutaneous cladribine on serum complement levels, which were also unchanged^47^.

The main limitation of this longitudinal study is the small number of participants, but this is offset by the frequency of blood sampling and lumbar punctures for CSF as well as the study duration. This study also lacks a control population (various formulations of cladribine are utilised in cancer), and therefore the study findings are not generalisable outside of MS.

In summary, this small but intensively monitored cohort study provides an in-depth perspective into the network relationships that exist in the immune system. Selective depletion in a certain cell population, in this case the memory B cells, leads to equilibration from mature plasmablasts and T cells, immature B cells and plasma cells. This is very different to that observed early on with alemtuzumab which has a more comprehensive depletory effect on T and B cells. The pharmacology of CladT is therefore very relevant to its selective targeting of B cells, particularly the memory B cells. Coinciding with this is a reduction in the B-cell chemoattractant CXCL-13 in the CSF and the IgG index. As a result, CladT influences intrathecal antibody production based on a reduction in кFLC, outside of its effect on oligoclonal bands. This possibly suggests that the impact on CSF immunoglobulin by CladT may be due to depletion of antibody secreting cell precursors in the periphery and CNS rather than by a major impact on plasmablasts and plasma cells that lose dCK during differentiation^9^. As such influence on immunoglobulins may take time. Since most of our current immunotherapies in MS particularly the monoclonal antibodies are not CNS penetrant this finding is very relevant to drug selection in RRMS.

## Data availability

Anonymised data are available from the authors upon reasonable request.

## Acknowledgments

The authors would like to thank all the participants in the study for their interest in the study and their contributions.

## Supplementary material

### Flow Cytometry and Panel Design

The panel was designed to incorporate a broad spectrum of commercially available antibodies, strategically selected for their ability to delineate diverse lymphocyte subsets (Table 1). The primary focus of the design was to delineate lymphocyte subsets, particularly emphasising T and B cells. Precision in the identification of these subsets was optimised through a strategic selection of fluorophore combinations. A notable advantage of utilizing the Cytek Biosciences 5-Laser Aurora system for full spectrum flow cytometry was its capacity to accommodate highly overlapping fluorochromes that traditionally posed compatibility challenges in conventional flow cytometers. Eighteen fluorochromes were chosen for this study, resulting in a panel configuration with the lowest complexity index (10.78), as illustrated in Supplementary Table 1 and Figure 1. In the process of fluorophore assignment to antigens, careful consideration was given to the brightness characteristics of each fluorophore. Particularly, brighter fluorophores were selectively assigned to stain low-density antigens, while dimmer fluorophores were purposefully chosen for high-intensity antigens, as detailed in Table 2. Pairs exhibiting the highest similarity indices, and consequently a greater predicted spread between them were only cFluor V420 and BV421 (0.97) but because these markers are not co-expressed there is no significant impact on the experiment. Due to the commercial availability and widespread use of all antigens in flow cytometry, antibody titration was excluded from the methodology. The recommended dilutions provided by the respective manufacturers were employed, ensuring adherence to established practices in the field. All results in the study are presented as absolute numbers and percentages of total lymphocytes. For the reference controls, beads were stained with the same fluorophores included in the panel and 19 single stainings were generated. The accuracy of the unmixing was tested by evaluating the unmixing results of the single cell staining compared to the result of a single tube containing all the fluorophores.

**Supplementary Table 1.**
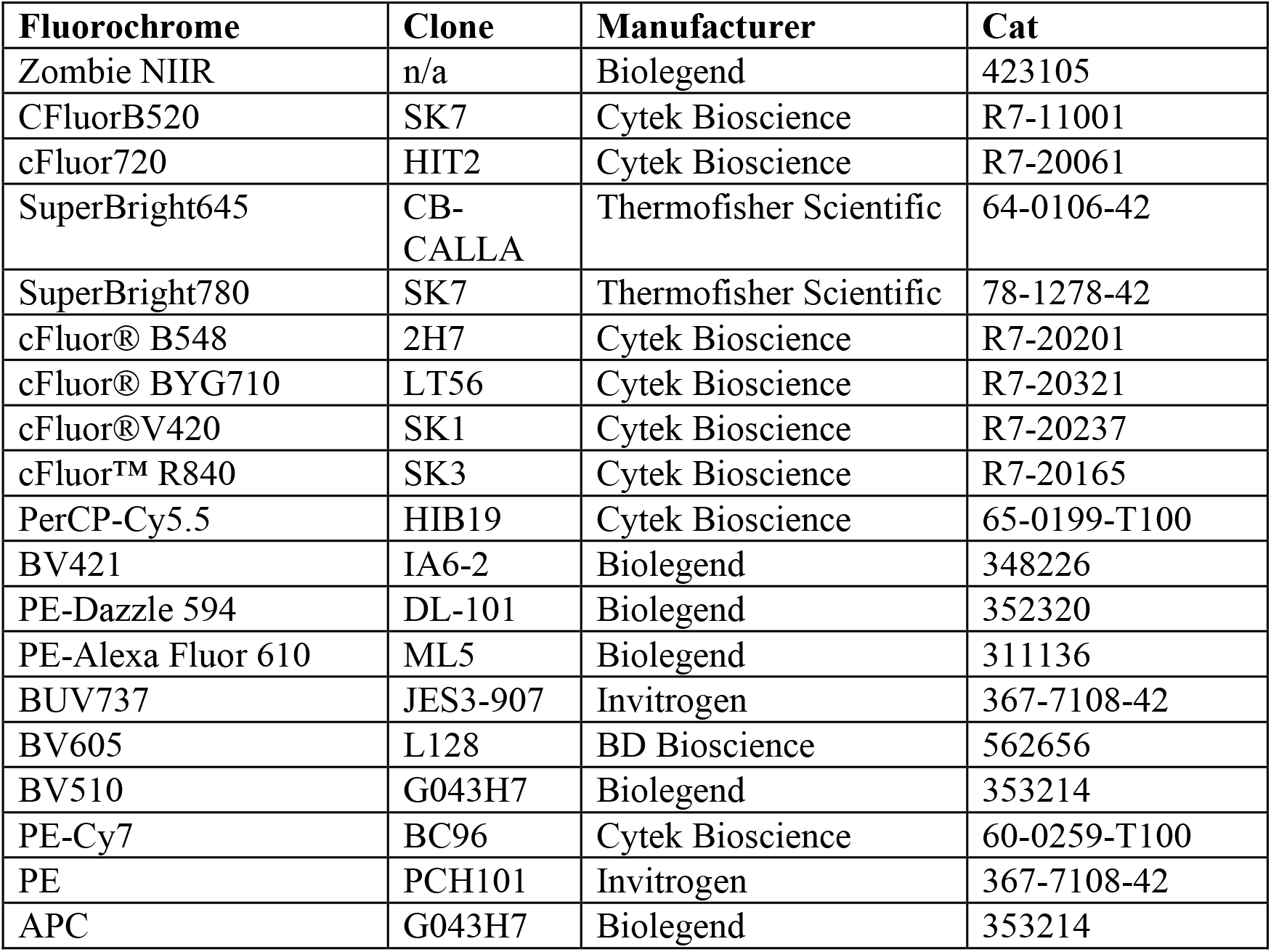
List of antibodies used in the study. Cell viability was determined using Live/ Dead Cell Stain, which is also included in the list.

**Supplementary Table 2.**
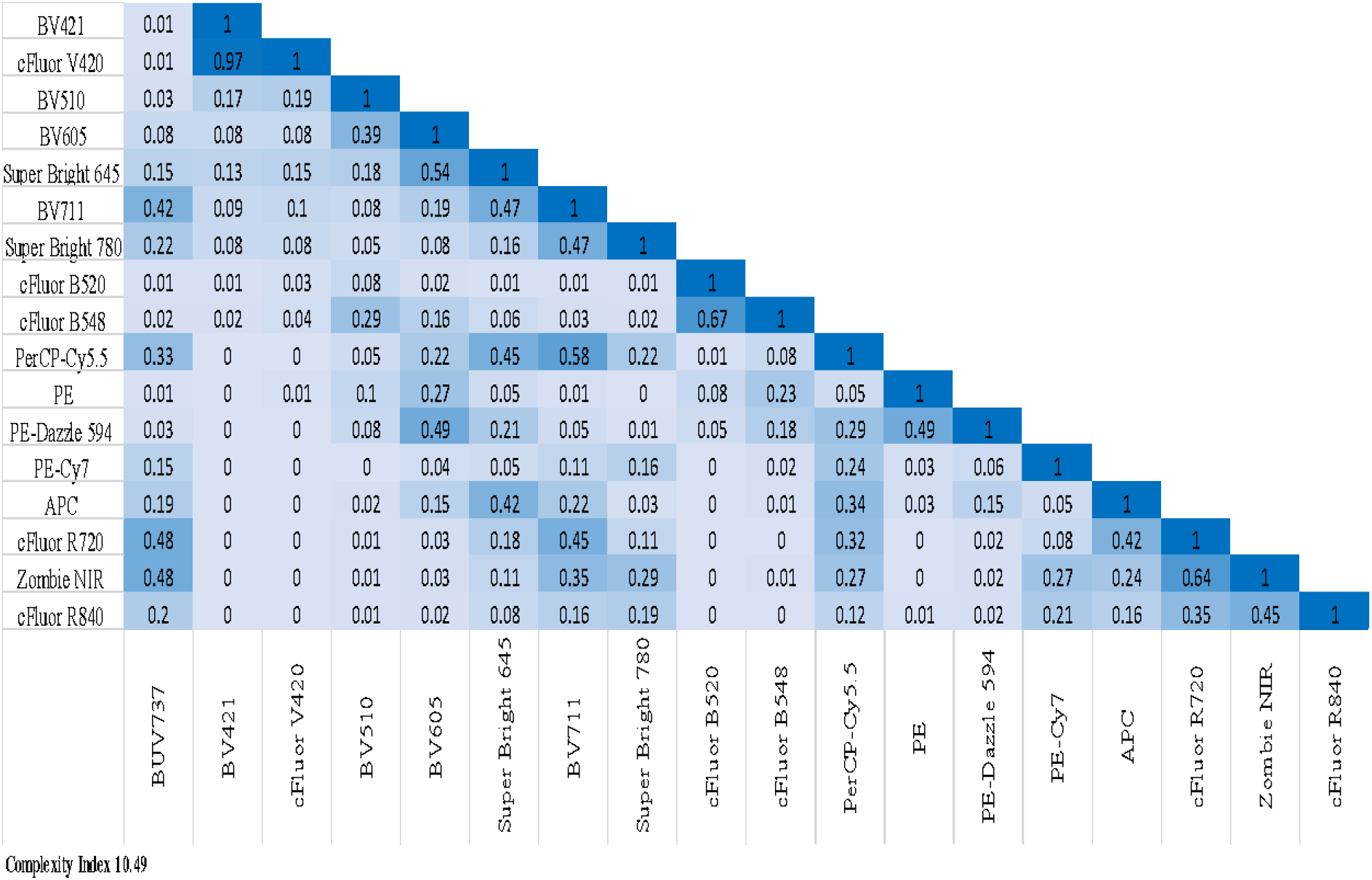
Similarity Index Matrix (SIM) of fluorochromes used in the panel.

**Supplementary Figure 1.**
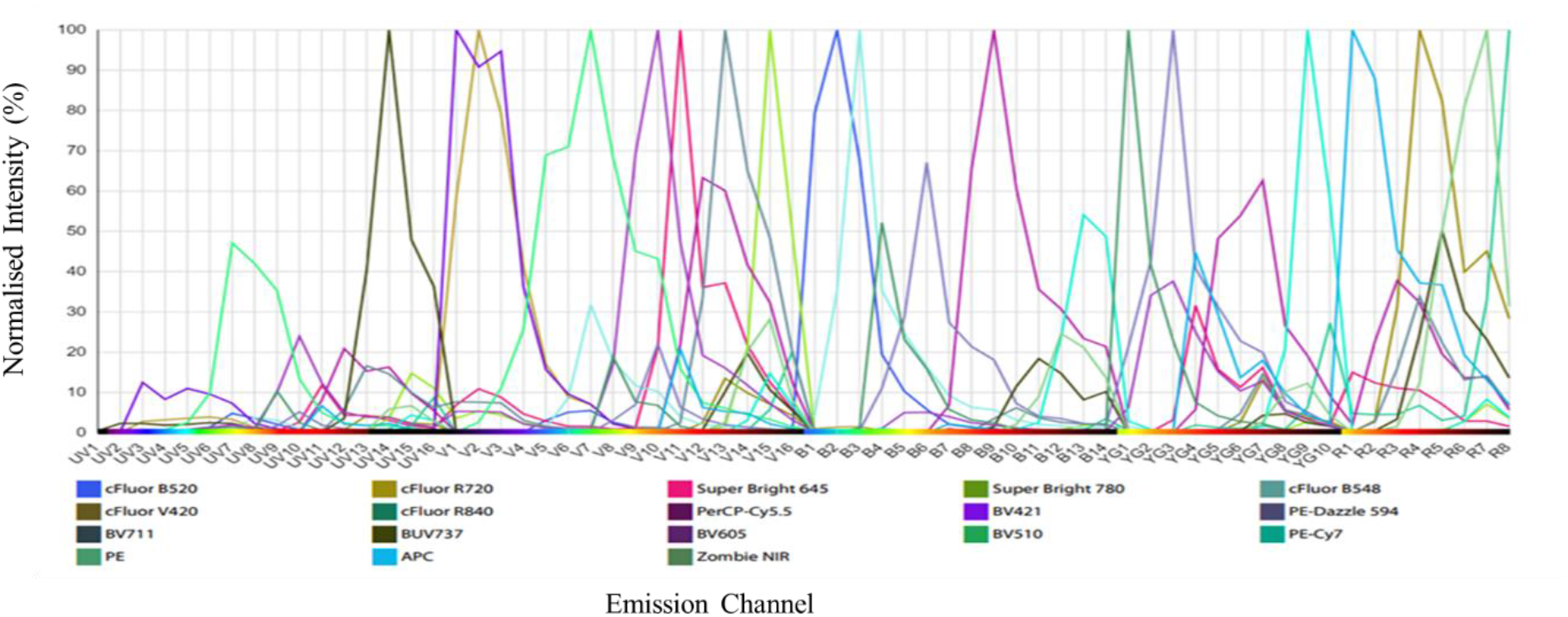
Full spectral analysis of the fluorophores used in the panel.

### Gating strategy

Initial gating includes the consideration of time to exclude events associated with technical anomalies. Further refinement involved the exclusion of doublets and aggregates using the parameters and side scatter (SSC)-A/H first and forward scatter (FSC) A/H. After isolation of live cells and exclusion of cellular debris, FSC and SSC were adjusted to optimally identify the lymphocyte population (Supplementary Figure 2).

For the T cell subsets, a comprehensive gating strategy was applied to the flow cytometry data (Supplementary Figure 3). From the lymphocytes gate, CD3 positive cells were initially selected for the T cell population (A). Within the CD3-positive gate, further discrimination was achieved to distinguish CD4^+^ and CD8^+^ T cell subsets, respectively (B). For characterization of T cell maturation both in CD4^+^ and CD8^+^ subsets, CCR7 and CD45RA were employed to distinguish between naïve, central memory (TCM), Effector memory (TEM) and terminally differentiated effector memory cells (TEMRA). Regulatory T cells (Tregs) were identified within the CD4^+^ helper cells as CD25^high^CD127low Foxp3^+^ (C).

For the B cell subsets (Supplementary Figure 4) initial gating involved the exclusion of T cells (CD3+). The CD19 marker was then used to identify the total B cell population within the lymphocyte gate. Further characterization of B cell subsets included gating on CD20 to refine the B cell identification. For the delineation of B cell maturation, CD27 and IgD were employed to distinguish between memory class switched, unswitched and naive B cells. Transitional B bells were obtained from Naïve cells. From CD19dimCD20^-^ gate, (B) CD38 and CD27 were utilised for additional refinement, aiding in the identification of plasmablast and providing insight into the B cell differentiation process. Further gates from CD19^dim^CD20 including CD27^-^IgD^-^ (C) were used to distinguish Plasma cells as CD138^+^CD38^+^. Within the CD19+ B cell population, regulatory cells were identified by gating on CD24^+^CD38^+^ cells (D).

**Supplementary Figure 2.**
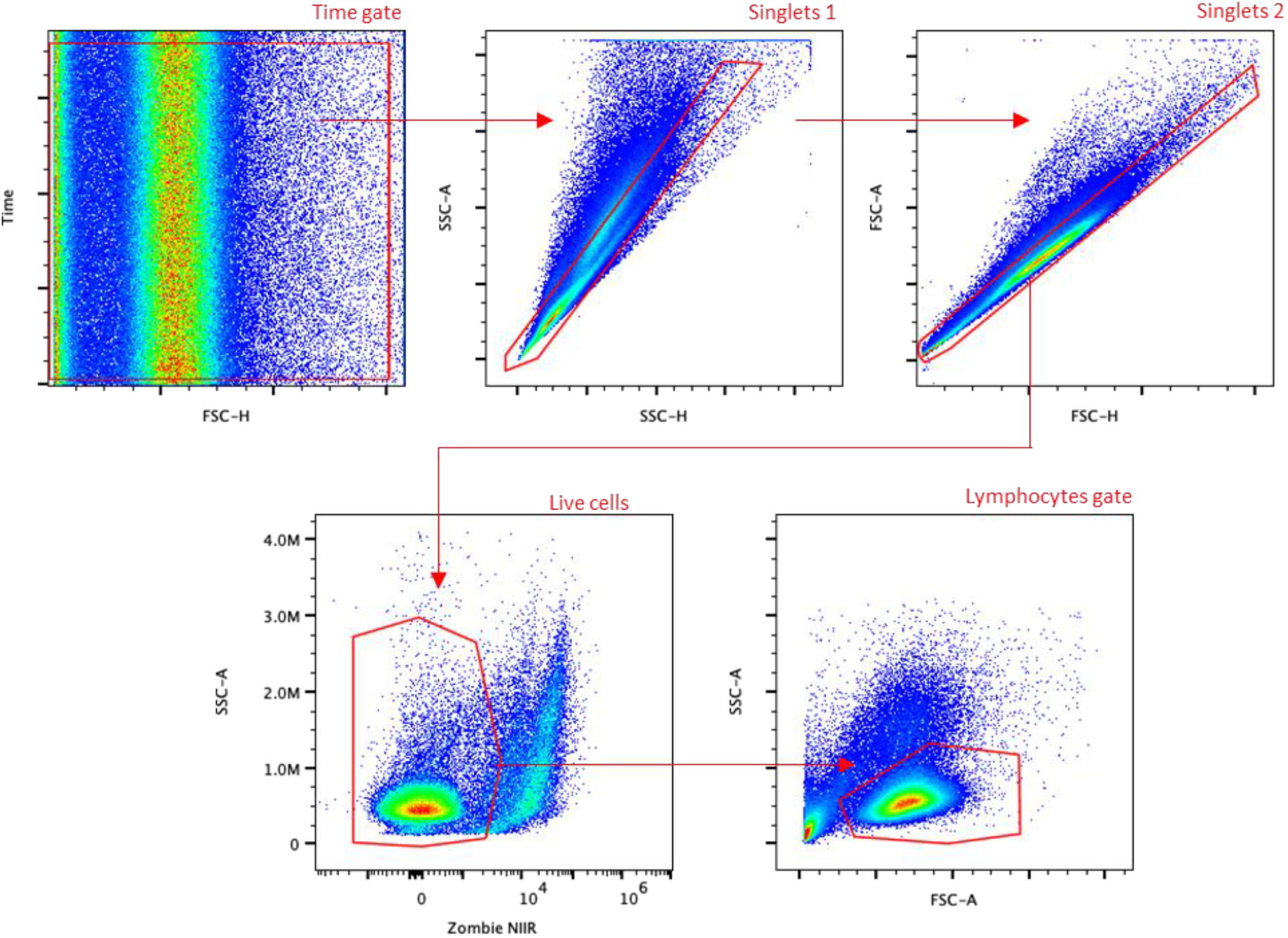
Spectral flow cytometry gating scheme to identify lymphocytes.

**Supplementary figure 3.**
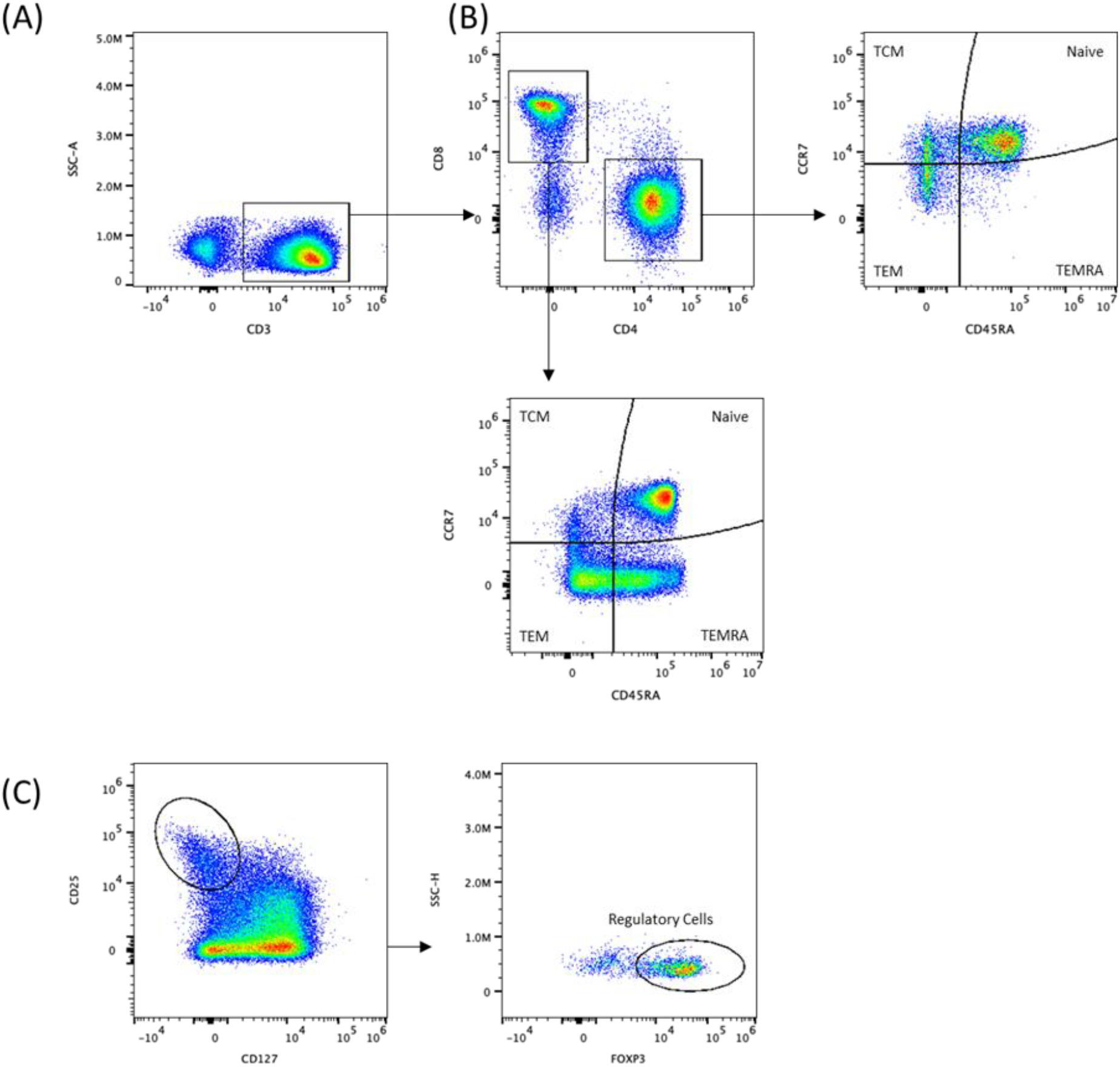
Flow cytometry gating scheme to identify T Lymphocytes.

**Supplementary figure 4.**
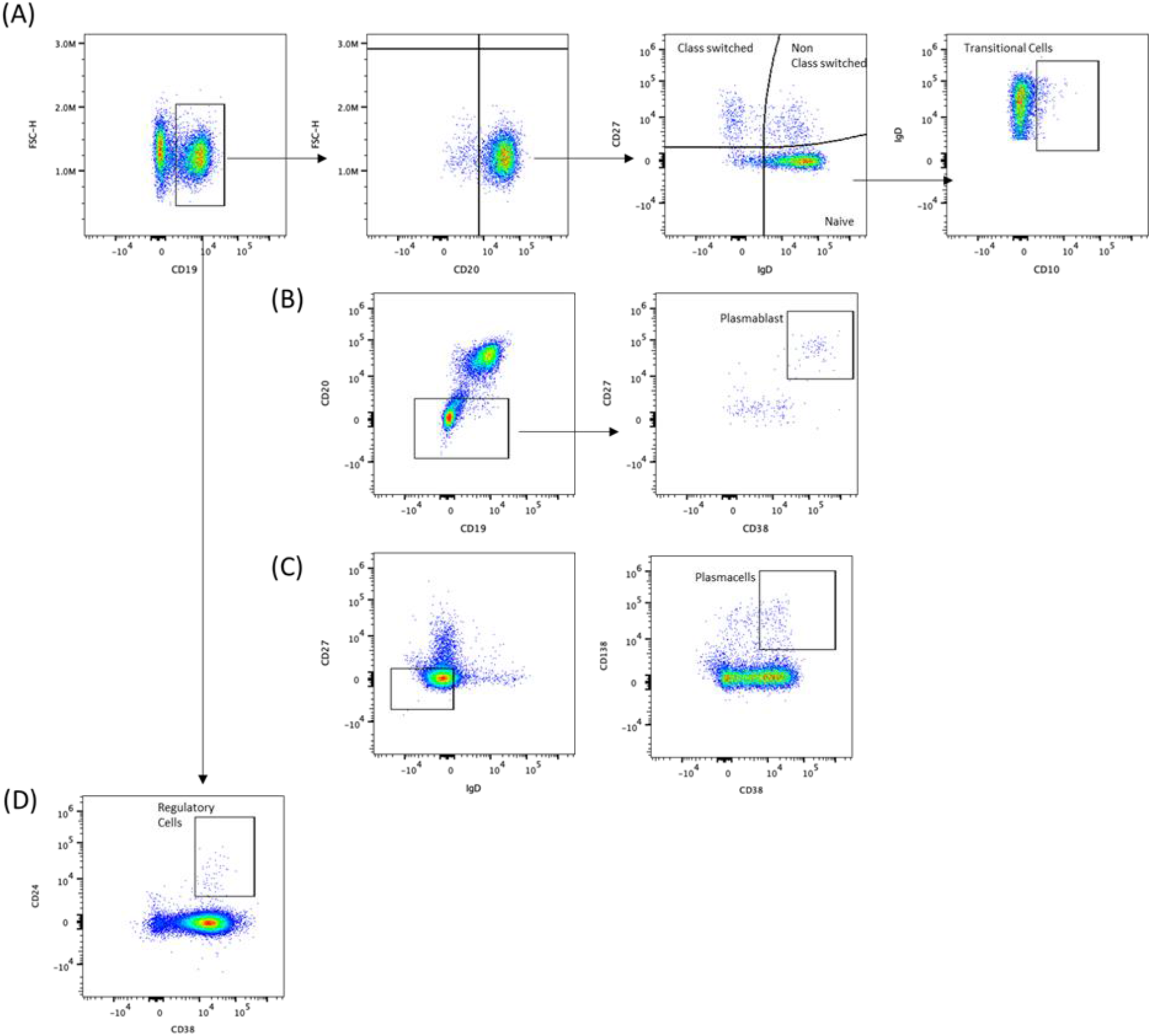
Flow cytometry gating scheme to identify B lymphocytes.

**Supplementary figure 5.**
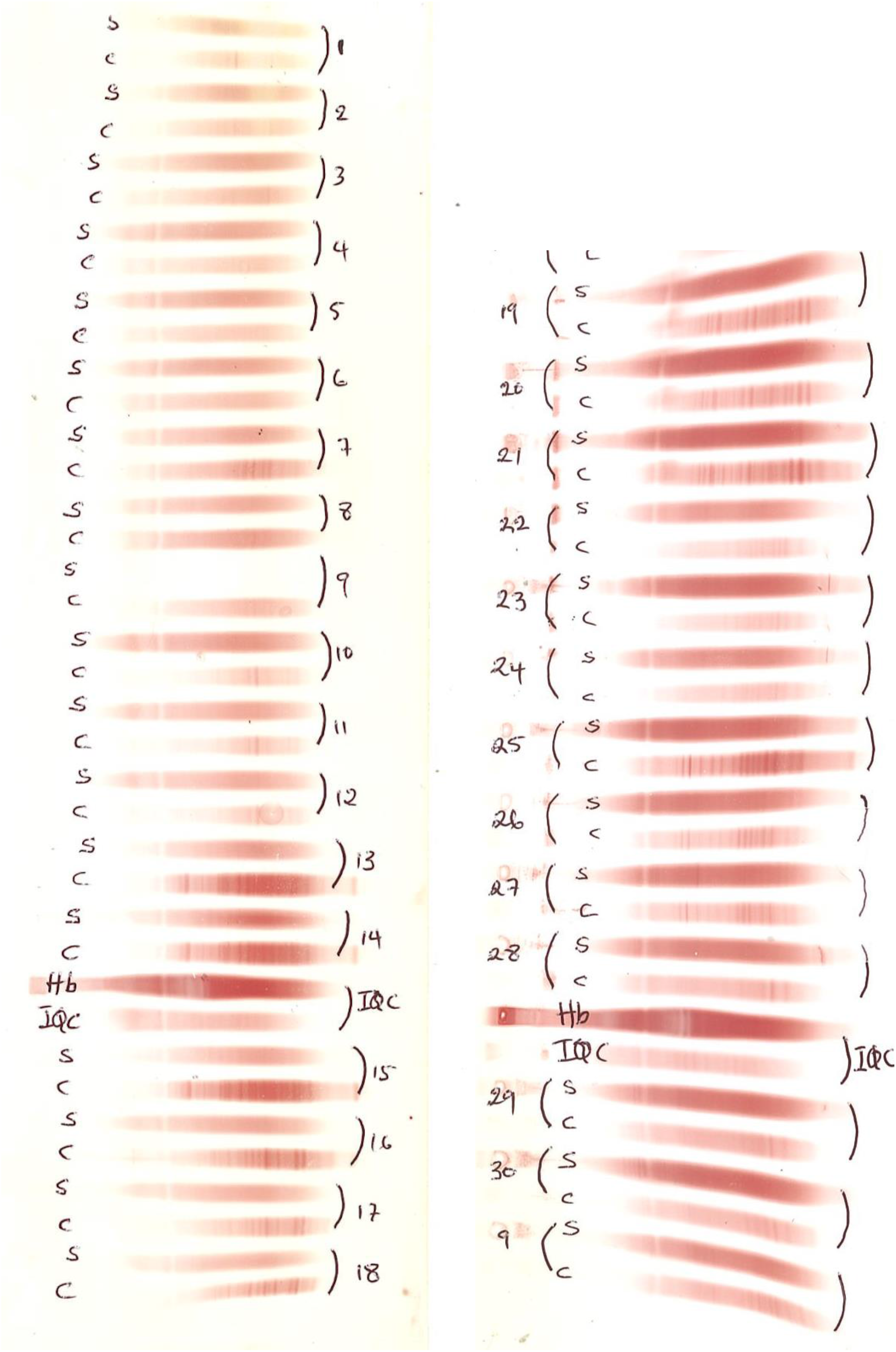
Oligoclonal bands (serum [s] and CSF [c], Hb and IQC quality controls)

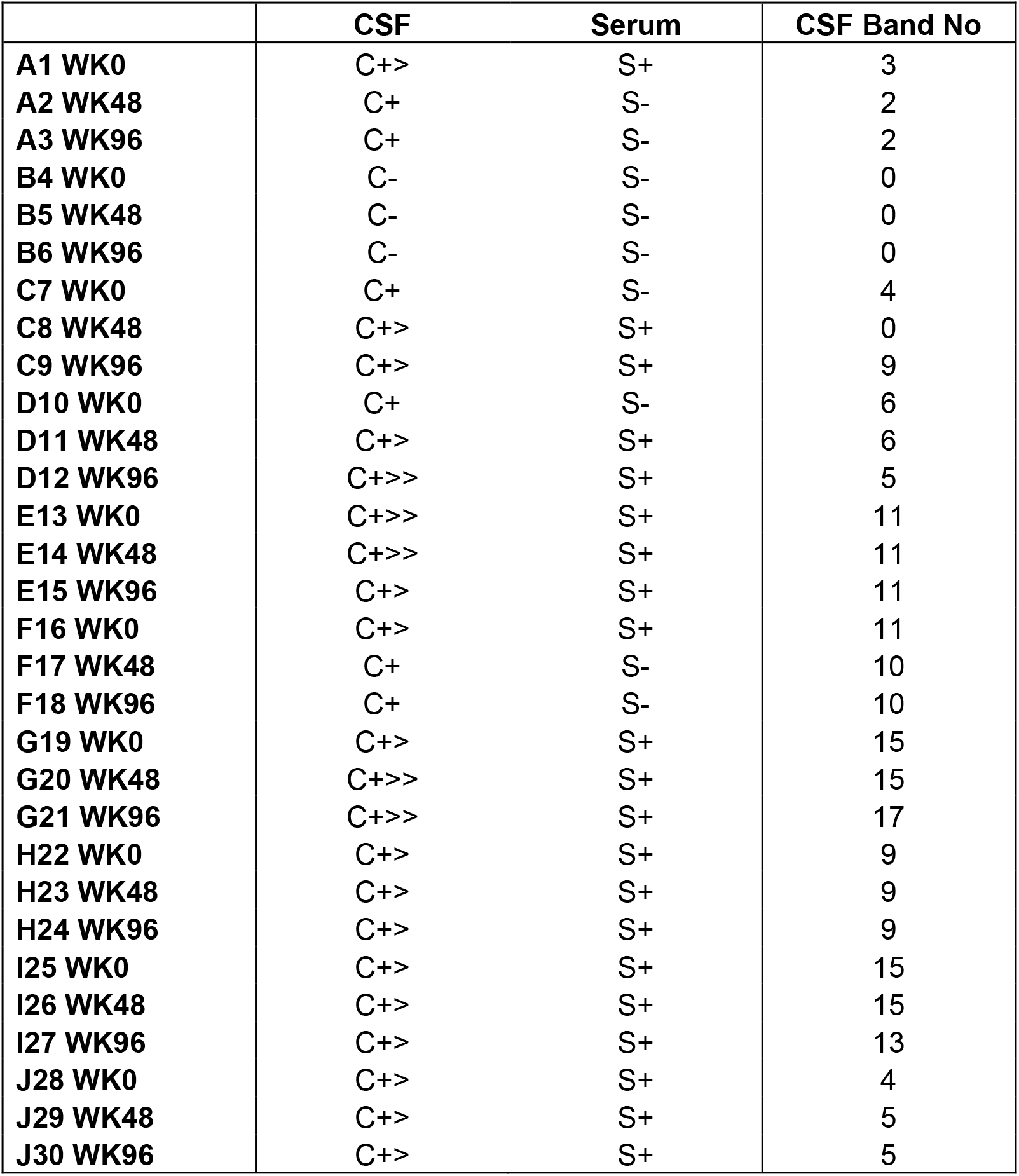

|  | **CSF** | **Serum** | **CSF Band No** |
| --- | --- | --- | --- |
| **A1 WK0** | C+> | S+ | 3 |
| **A2 WK48** | C+ | S- | 2 |
| **A3 WK96** | C+ | S- | 2 |
| **B4 WK0** | C- | S- | 0 |
| **B5 WK48** | C- | S- | 0 |
| **B6 WK96** | C- | S- | 0 |
| **C7 WK0** | C+ | S- | 4 |
| **C8 WK48** | C+> | S+ | 0 |
| **C9 WK96** | C+> | S+ | 9 |
| **D10 WK0** | C+ | S- | 6 |
| **D11 WK48** | C+> | S+ | 6 |
| **D12 WK96** | C+>> | S+ | 5 |
| **E13 WK0** | C+>> | S+ | 11 |
| **E14 WK48** | C+>> | S+ | 11 |
| **E15 WK96** | C+> | S+ | 11 |
| **F16 WK0** | C+> | S+ | 11 |
| **F17 WK48** | C+ | S- | 10 |
| **F18 WK96** | C+ | S- | 10 |
| **G19 WK0** | C+> | S+ | 15 |
| **G20 WK48** | C+>> | S+ | 15 |
| **G21 WK96** | C+>> | S+ | 17 |
| **H22 WK0** | C+> | S+ | 9 |
| **H23 WK48** | C+> | S+ | 9 |
| **H24 WK96** | C+> | S+ | 9 |
| **I25 WK0** | C+> | S+ | 15 |
| **I26 WK48** | C+> | S+ | 15 |
| **I27 WK96** | C+> | S+ | 13 |
| **J28 WK0** | C+> | S+ | 4 |
| **J29 WK48** | C+> | S+ | 5 |
| **J30 WK96** | C+> | S+ | 5 |

**Supplementary Table 3.** Biomarker results (mean, standard deviation, median)

|  | Baseline |  |  | Week 48 |  |  | Week 96 |  |  |
| --- | --- | --- | --- | --- | --- | --- | --- | --- | --- |
|  | Mean | SD | Median | Mean | SD | Median | Mean | SD | Median |
| **KFL Index** | 164.52 | 227.06 | 61.29 | 71.31 | 84.69 | 43.47 | 64.40 | 67.25 | 37.84 |
| **LFL Index** | 31.44 | 26.99 | 24.93 | 27.88 | 25.23 | 20.13 | 27.58 | 28.66 | 17.49 |
| **IgG Index** | 1.15 | 0.52 | 0.93 | 0.83 | 0.35 | 0.77 | 0.82 | 0.35 | 0.63 |
| **IgA index** | 0.21 | 0.04 | 0.21 | 0.24 | 0.13 | 0.22 | 0.21 | 0.04 | 0.23 |
| **IgM index** | 0.20 | 0.31 | 0.07 | 0.08 | 0.06 | 0.07 | 0.07 | 0.06 | 0.05 |
| **CSF Complement C3** | 1.14 | 0.17 | 1.12 | 1.16 | 0.17 | 1.10 | 1.19 | 0.22 | 1.15 |
| **CSF IFNγ** | 1.09 | 0.75 | 0.89 | 0.99 | 0.90 | 0.66 | 1.01 | 1.22 | 0.63 |
| **CSF IL10** | 0.11 | 0.11 | 0.08 | 0.13 | 0.19 | 0.05 | 0.06 | 0.04 | 0.06 |
| **CSF IL12p70** | 0.02 | 0.03 | 0.00 | 0.01 | 0.01 | 0.01 | 0.02 | 0.03 | 0.00 |
| **CSF IL13** | 0.23 | 0.27 | 0.17 | 0.22 | 0.24 | 0.09 | 0.31 | 0.20 | 0.38 |
| **CSF IL1b** | 0.11 | 0.05 | 0.12 | 0.11 | 0.05 | 0.11 | 0.08 | 0.04 | 0.06 |
| **CSF IL2** | 0.02 | 0.03 | 0.01 | 0.02 | 0.03 | 0.01 | 0.02 | 0.02 | 0.01 |
| **CSF IL4** | 0.00 | 0.00 | 0.00 | 0.00 | 0.00 | 0.00 | 0.00 | 0.01 | 0.00 |
| **CSF IL5** | 0.18 | 0.09 | 0.18 | 0.25 | 0.13 | 0.28 | 0.21 | 0.14 | 0.16 |
| **CSF IL8** | 27.11 | 13.05 | 27.79 | 29.65 | 9.56 | 28.32 | 25.56 | 12.74 | 21.62 |
| **CSF TNFα** | 0.05 | 0.04 | 0.03 | 0.04 | 0.05 | 0.03 | 0.06 | 0.06 | 0.04 |
| **CSF NfL** | 869.27 | 1096.75 | 534.42 | 287.65 | 90.01 | 271.77 | 502.93 | 721.95 | 301.13 |
| **CSF CXCL13** | 88.55 | 68.45 | 73.12 | 39.43 | 35.16 | 34.53 | 19.10 | 11.67 | 13.88 |
| **CSF sCD138** | 0.25 | 0.28 | 0.11 | 0.14 | 0.20 | 0.05 | 0.09 | 0.09 | 0.05 |
| **Urine Neopterin** | 224.64 | 268.06 | 109.15 | 355.87 | 566.27 | 92.17 | 136.99 | 114.18 | 112.70 |

**Supplementary table 4.** Spearman Correlation between Baseline, 48wh and 96wks in T cells (p<0.05 in bold)

|  |  | % Change BL to 48 wks |  | % Change BL to 96 wks |  |
| --- | --- | --- | --- | --- | --- |
|  | Outcome | r | p-value | r | p-value |
| **T-cells** |  |  |  |  |  |
| **CD3<sup>+</sup>** | EDSS | 0.102 | 0.791 | 0.120 | 0.771 |
|  | PASAT | -0.024 | 0.954 | -0.429 | 0.330 |
|  | MSIS29 | -0.100 | 0.794 | 0.468 | 0.283 |
|  | PEGDH | 0.619 | 0.102 | -0.036 | 0.938 |
|  | PEGnonDH | 0.119 | 0.775 | 0.357 | 0.424 |
|  | FWTrial | 0.000 | 1.000 | -0.257 | 0.615 |
|  | T2 lesions | 0.427 | 0.217 | 0.320 | 0.377 |
|  | Gd lesions | 0.290 | 0.502 | 0.406 | 0.299 |
| **CD3<sup>+</sup>CD4<sup>+</sup>** | EDSS | 0.179 | 0.641 | 0.072 | 0.861 |
|  | PASAT | -0.048 | 0.909 | -0.429 | 0.330 |
|  | MSIS29 | -0.033 | 0.931 | 0.468 | 0.283 |
|  | PEGDH | 0.595 | 0.119 | -0.036 | 0.938 |
|  | PEGnonDH | 0.190 | 0.645 | 0.357 | 0.424 |
|  | FWTrial | 0.000 | 1.000 | -0.257 | 0.615 |
|  | T2 lesions | 0.360 | 0.305 | 0.320 | 0.377 |
|  | Gd lesions | 0.290 | 0.502 | 0.406 | 0.299 |
| **CD3<sup>+</sup>CD8<sup>+</sup>** | EDSS | 0.077 | 0.842 | 0.554 | 0.152 |
|  | PASAT | -0.048 | 0.909 | -0.821 | **0.029** |
|  | MSIS29 | -0.033 | 0.931 | 0.559 | 0.190 |
|  | PEGDH | 0.714 | **0.050** | 0.214 | 0.637 |
|  | PEGnonDH | 0.262 | 0.524 | 0.714 | 0.074 |
|  | FWTrial | 0.095 | 0.819 | 0.257 | 0.615 |
|  | T2 lesions | 0.674 | **0.034** | 0.372 | 0.298 |
|  | Gd lesions | 0.522 | 0.090 | 0.058 | 0.901 |
|  | PEGnonDH | 0.357 | 0.378 | 0.143 | 0.755 |
|  | FWTrial | -0.048 | 0.909 | -0.143 | 0.782 |
|  | T2 lesions | 0.360 | 0.305 | 0.415 | 0.239 |
|  | Gd lesions | 0.290 | 0.502 | 0.406 | 0.299 |

**Supplementary table 4 continued**
|  |  | % Change BL to 48 wks |  | % Change BL to 96 wks |  |
| --- | --- | --- | --- | --- | --- |
|  | **Outcome** | **r** | **p-value** | **r** | **p-value** |
| **T-cells** |  |  |  |  |  |
| **CD4 Tcm** | EDSS | -0.060 | 0.877 | 0.072 | 0.861 |
|  | PASAT | -0.024 | 0.954 | -0.714 | 0.074 |
|  | MSIS29 | -0.133 | 0.728 | 0.414 | 0.349 |
|  | PEGDH | 0.548 | 0.158 | -0.179 | 0.695 |
|  | PEGnonDH | 0.143 | 0.731 | 0.357 | 0.424 |
|  | FWTrial | -0.048 | 0.909 | -0.143 | 0.782 |
|  | T2 lesions | 0.494 | 0.146 | 0.493 | 0.150 |
|  | Gd lesions | 0.290 | 0.502 | 0.522 | 0.090 |
| **CD4 Naïve** | EDSS | 0.289 | 0.444 | 0.036 | 0.930 |
|  | PASAT | -0.167 | 0.688 | -0.464 | 0.288 |
|  | MSIS29 | 0.050 | 0.896 | 0.378 | 0.396 |
|  | PEGDH | 0.810 | **0.019** | -0.429 | 0.330 |
|  | PEGnonDH | 0.310 | 0.448 | 0.036 | 0.938 |
|  | FWTrial | 0.190 | 0.645 | -0.486 | 0.321 |
|  | T2 lesions | 0.360 | 0.305 | 0.320 | 0.377 |
|  | Gd lesions | 0.290 | 0.502 | 0.406 | 0.299 |
| **CD4 Temra** | EDSS | 0.170 | 0.657 | 0.217 | 0.597 |
|  | PASAT | -0.548 | 0.158 | -0.714 | 0.074 |
|  | MSIS29 | 0.050 | 0.896 | 0.505 | 0.244 |
|  | PEGDH | 0.857 | **0.010** | -0.286 | 0.526 |
|  | PEGnonDH | 0.357 | 0.378 | 0.143 | 0.755 |
|  | FWTrial | -0.048 | 0.909 | -0.143 | 0.782 |
|  | T2 lesions | 0.360 | 0.305 | 0.415 | 0.239 |
|  | Gd lesions | 0.290 | 0.502 | 0.406 | 0.299 |
| **CD4 Tem** | EDSS | 0.179 | 0.641 | 0.048 | 0.907 |
|  | PASAT | -0.024 | 0.954 | -0.321 | 0.474 |
|  | MSIS29 | -0.150 | 0.695 | 0.162 | 0.723 |
|  | PEGDH | 0.619 | 0.102 | -0.321 | 0.474 |
|  | PEGnonDH | 0.214 | 0.604 | 0.143 | 0.755 |
|  | FWTrial | -0.119 | 0.775 | -0.600 | 0.204 |
|  | T2 lesions | 0.517 | 0.126 | 0.415 | 0.239 |
|  | Gd lesions | 0.406 | 0.299 | 0.406 | 0.299 |

**Supplementary table 4 continued**
|  |  | % Change BL to 48 wks |  | % Change BL to 96 wks |  |
| --- | --- | --- | --- | --- | --- |
|  | Outcome | r | p-value | r | p-value |
| **T-cells** |  |  |  |  |  |
| **CD8 Tcm** | EDSS | 0.153 | 0.690 | 0.422 | 0.291 |
|  | PASAT | 0.048 | 0.909 | -0.393 | 0.376 |
|  | MSIS29 | -0.067 | 0.862 | 0.252 | 0.578 |
|  | PEGDH | 0.690 | 0.061 | 0.321 | 0.474 |
|  | PEGnonDH | 0.333 | 0.412 | 0.679 | 0.095 |
|  | FWTrial | 0.095 | 0.819 | 0.029 | 0.956 |
|  | T2 lesions | 0.607 | 0.064 | 0.372 | 0.298 |
|  | Gd lesions | 0.522 | 0.090 | 0.058 | 0.901 |
| **CD8 Naïve** | EDSS | 0.247 | 0.516 | 0.506 | 0.196 |
|  | PASAT | -0.167 | 0.688 | -0.821 | **0.029** |
|  | MSIS29 | 0.083 | 0.828 | 0.559 | 0.190 |
|  | PEGDH | 0.833 | **0.014** | 0.214 | 0.637 |
|  | PEGnonDH | 0.381 | 0.345 | 0.714 | 0.074 |
|  | FWTrial | 0.167 | 0.688 | 0.257 | 0.615 |
|  | T2 lesions | 0.607 | 0.064 | 0.372 | 0.298 |
|  | Gd lesions | 0.522 | 0.090 | 0.058 | 0.901 |
| **CD8 Temra** | EDSS | 0.128 | 0.740 | -0.108 | 0.793 |
|  | PASAT | 0.071 | 0.864 | -0.357 | 0.424 |
|  | MSIS29 | -0.350 | 0.350 | -0.018 | 0.969 |
|  | PEGDH | 0.595 | 0.119 | -0.429 | 0.330 |
|  | PEGnonDH | 0.048 | 0.909 | 0.036 | 0.938 |
|  | FWTrial | -0.048 | 0.909 | -0.600 | 0.204 |
|  | T2 lesions | 0.247 | 0.488 | 0.320 | 0.377 |
|  | Gd lesions | 0.058 | 0.901 | 0.406 | 0.299 |
| **CD8 Tcm** | EDSS | -0.017 | 0.965 | 0.313 | 0.440 |
|  | PASAT | 0.000 | 1.000 | 0.036 | 0.938 |
|  | MSIS29 | -0.267 | 0.481 | 0.252 | 0.578 |
|  | PEGDH | 0.619 | 0.102 | 0.536 | 0.211 |
|  | PEGnonDH | 0.119 | 0.775 | 0.643 | 0.120 |
|  | FWTrial | -0.119 | 0.775 | -0.086 | 0.868 |
|  | T2 lesions | 0.652 | **0.043** | 0.121 | 0.746 |
|  | Gd lesions | 0.406 | 0.299 | -0.174 | 0.702 |
| **Regulatory Cells** | EDSS | 0.196 | 0.609 | 0.506 | 0.196 |
|  | PASAT | -0.048 | 0.909 | -0.107 | 0.815 |
|  | MSIS29 | 0.017 | 0.965 | 0.559 | 0.190 |
|  | PEGDH | 0.738 | **0.040** | 0.714 | 0.074 |
|  | PEGnonDH | 0.286 | 0.485 | 0.750 | 0.056 |
|  | FWTrial | 0.095 | 0.819 | 0.200 | 0.697 |
|  | T2 lesions | 0.607 | 0.064 | 0.277 | 0.449 |
|  | Gd lesions | 0.522 | 0.090 | 0.058 | 0.901 |

**Supplementary table 5.**
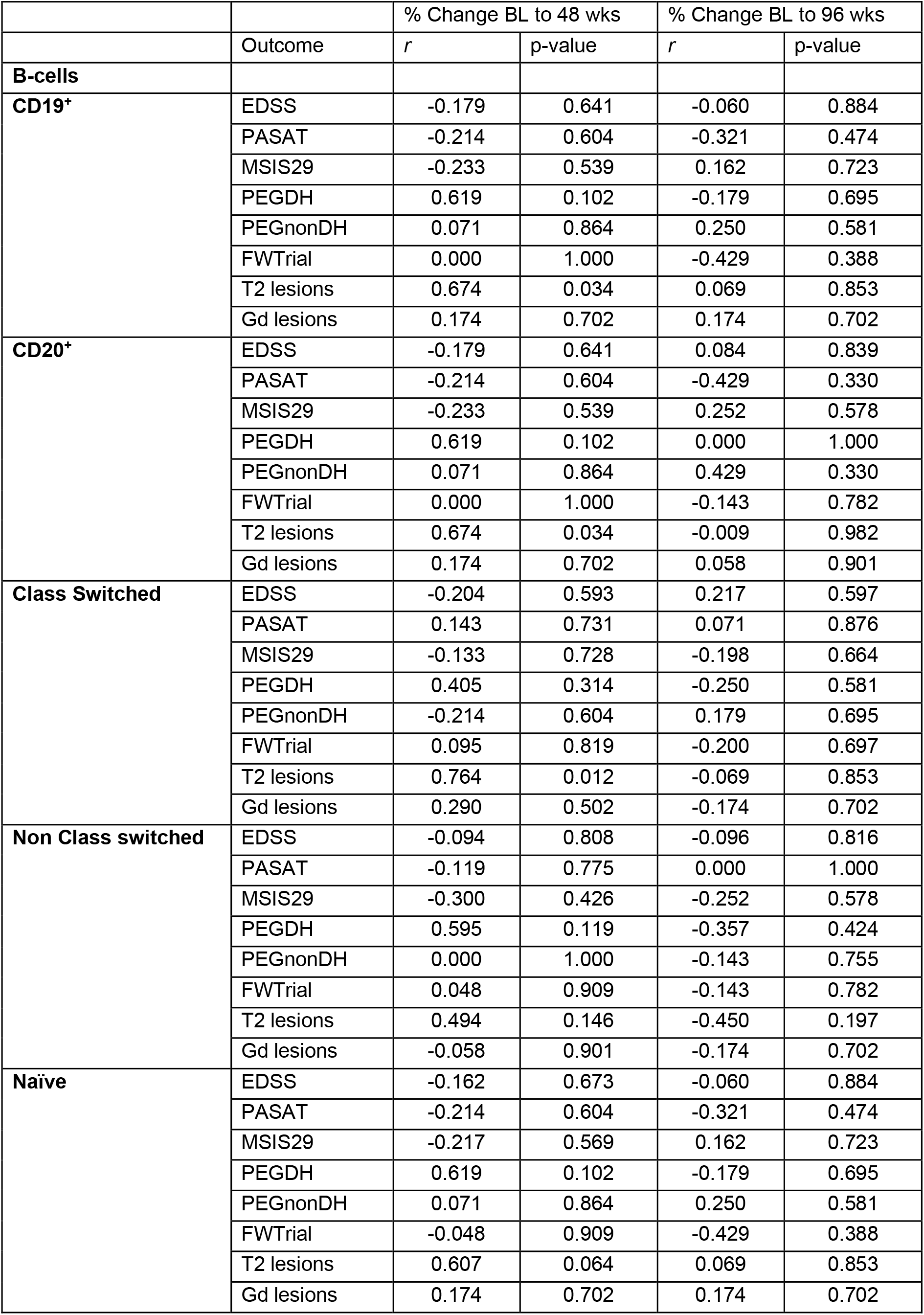
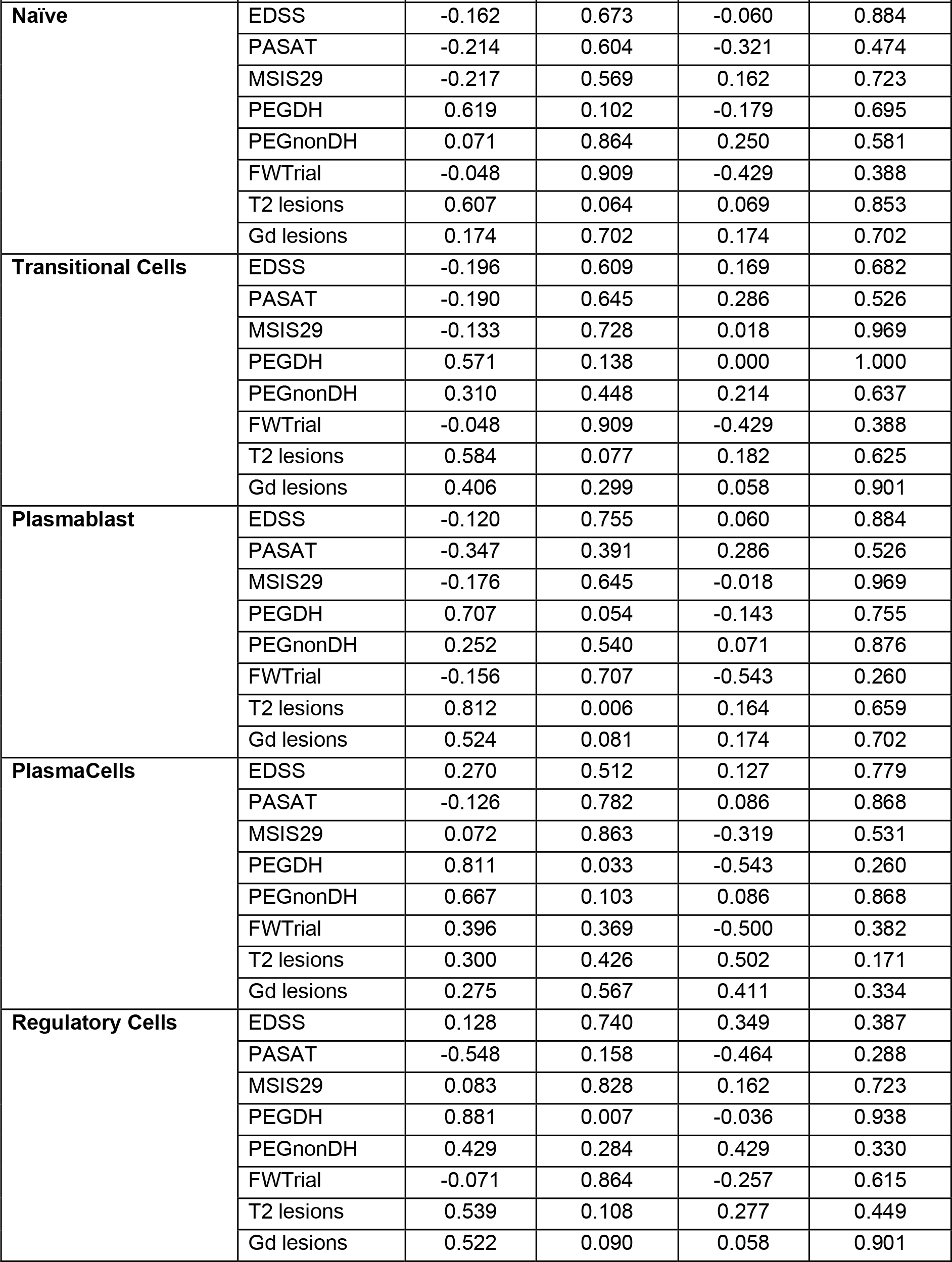
Spearman Correlation between Baseline, 48wh and 96wks in B cells.

